# Preliminary Validation of a Multidimensional Stimulability Testing Rating Scale for Patients with Voice Disorders

**DOI:** 10.64898/2026.10.13.26365451

**Authors:** Elizabeth D. Young, Laura E. Toles

**Affiliations:** School of Behavioral and Brain Sciences, University of Texas at Dallas, Richardson, TX; Department of Otolaryngology-Head and Neck Surgery, University of Texas Southwestern Medical Center

**Keywords:** Stimulability testing, Voice assessment, Dysphonia

## Abstract

**Purpose:** Stimulability testing involves the evaluation of multiple aspects of a patient’s behavior, but the results are most frequently consolidated into a single domain (i.e., “stimulable”). The current study performs preliminary content, construct, concurrent, and predictive validation for the Multidimensional Stimulability Profile for Voice (MSP-V), a rating scale designed to measure multiple domains during stimulability testing (Degree of Change, Rapidity of Change, Generalization, Patient-rated Voice Quality, Patient-rated Voice Feel, and an overall rating of Stimulability Level).

**Methods:** 78 patients with voice disorders (24 functional, 15 structural, 39 neurological) were administered the MSP-V by five speech-language pathologists (SLPs). Content validity was evaluated via the Content Validity Index (CVI) completed by nine voice-specialized SLPs. Construct validity was evaluated via exploratory factor analysis. Concurrent validity was assessed by determining whether MSP-V domains scaled in the expected direction compared to categorical stimulability ratings (e.g., “Highly stimulable” vs. Stimulable”). Predictive validity was assessed by examining the relationship between MSP-V domains and voice disorder diagnosis, return to voice therapy, and patient-reported practice outside of therapy.

**Results:** The CVI reached the critical value for clinician-rated and patient-rated domains. Two distinct factors were identified during EFA, one consisting of clinician-rated domains and the other of patient-rated domains. All MSP-V domains showed the expected directional relationships as categorical stimulability ratings. No domains differed significantly between disorder etiologies. The Rapidity of Change and Stimulability Level domains both significantly predicted return for voice therapy, where participants who rated lower on each domain were more likely to return to for voice therapy. No domains were predictive of patient-reported practice.

**Conclusions:** The MSP-V is a clinically feasible tool with emerging content, construct, concurrent, and predictive validity that could be used to provide increased clarity and precision in stimulability testing documentation for research and practice.

## Introduction

Stimulability testing, the evaluation of an individual’s ability to modify a behavior using therapeutic techniques, is a highly-valued part of the behavioral voice evaluation (Coleman et al., 2025; Gillespie & Gartner-Schmidt, 2016; Petty et al., 2023; Toles et al., 2024; Toles & Young, 2023). Both laryngologists and voice-specialized speech-language pathologists (SLPs) report performing stimulability testing (ST) with their voice patients to inform clinical decisions such as determining patient candidacy for behavioral intervention, differential diagnosis, and estimating prognosis for improvement with behavioral therapy (Coleman et al., 2025; Toles & Young, 2023). However, ST practices are not widely standardized; less than 15% of laryngologists and less than 10% of SLPs reported using a standard ST procedure in their current practice setting. As a result, there is currently no validated standardized method of measuring ST outcomes.

ST outcomes are often summarized using the term “stimulable” (e.g., McDowell et al., 2023; Shelly et al., 2023; Toles et al., 2024; Weston et al., 2023). For example, a person or behavior may be “stimulable”, “somewhat stimulable”, or “not stimulable” based on their response to ST. However, practicing voice-specialized SLPs report a variety of underlying domains contribute to their evaluation of this single outcome. Young (2025) interviewed eight voice-specialized SLPs about their definitions of what it meant to be “stimulable” before and after performing mock ST trials on individuals without voice complaints. The eight SLPs described twenty-seven domains that they considered when assessing the extent to which a person or behavior is stimulable. These domains included the degree of behavioral change achieved, the patient’s ability to transfer or maintain behavioral change, the degree of clinician support required by the patient during ST, and psychosocial factors influencing the patient’s ability and motivation to participate in ST. All clinicians described evaluating multiple domains during ST; however, only one domain out of 27 – rapidity of voice change – was discussed by all eight SLPs. Additionally, five of the 27 domains (19%) only emerged in discussion following mock ST trials. These findings, while preliminary, suggest a lack of a common understanding of what is being evaluated during ST, as well as a mismatch between the conceptualization and actualization of measuring ST outcomes.

The pattern found in Young (2025) is mirrored in the literature regarding ST. Studies that describe ST outcomes frequently do so without adequate detail for reproduction. For example, two publications describe measuring ST outcomes using the following criteria: “performance on a cued task is better than the original performance on the same task” (Gillespie & Gartner-Schmidt, 2016; Shelly et al., 2023). This definition suggests performance on the task must be “better” but does not describe how “better” is measured (i.e., a positive change in clinician-perceived voice quality vs. positive change in patient-perceived voice quality vs. positive change in acoustic parameters, etc.). Other studies will specify multiple domains examined during ST (e.g., “the voice was described as clearer, more efficient, or otherwise enhanced”, “aware[ness] of acute behavioral changes”, “make behavioral changes with minimal cueing”), but collapse all domains into the single-domain “stimulable” vs “not-stimulable”, (e.g., McDowell et al., 2023; Turner & Toles, 2025). Additionally, many articles specify broad outcomes of ST that can be either individual-specific (e.g., “stimulable for voice therapy”; Chen et al., 2020; Gates et al., 2024; Grillo & Wolfberg, 2023; McDowell et al., 2023), or behavior-specific (e.g., “stimulable for resonant voice” De Almeida et al., 2019; Gartner-Schmidt & Gillespie, 2021; Kempster et al., 2009). While this wording specifies the general target of ST, it does not provide information regarding the underlying mechanisms that are being measured during such testing.

As a result of the lack of standardization in ST practices and reporting, ST outcomes are poorly described and understood. The pattern within the literature suggests that clinicians likely measure multiple domains during ST when determining whether an individual or behavior is “stimulable”. However, because there is currently no method of measuring these underlying domains, it is unknown which domains contribute to success of ST. Because of this, clinicians currently do not have strong scientific evidence to draw upon when conducting ST and are instead forced to rely on clinical intuition. The lack of standardization also introduces significant clinician bias when making clinical decisions and communicating with other voice care professionals. As a result, the potential predictive power of ST is greatly reduced, resulting in increased cost and burden of care.

The Multidimensional Stimulability Profile for Voice (MSP-V) was designed to provide a standardized method of evaluating multiple domains of ST. The MSP-V contains six domains: Degree of Change, Rapidity of Change, Generalization, Stimulability Level, Patient-Rated Change in Voice Sound, and Patient-Rated Change in Voice Feel. Each domain, which will be described in depth below, were selected by a combination of expert opinion, clinical feasibility testing, and review of the available literature. The MSP-V was additionally designed to provide clinicians with an easy, accessible method of rating ST outcomes within a clinical context regardless of their personal ST style.

The goal of the current study was to gather preliminary information regarding the validity of the MSP-V to inform its continued development as a rating scale for ST. To do this, initial examination of the content, construct, concurrent, and predictive validity of the MSP-V was performed using pilot data collected for ST performed on 78 patients at a single clinical site. Validating a measure that quantifies the results of ST will provide needed clarity and precision when evaluating ST outcomes in research and in practice. Further, using a multidomain metric for quantifying ST results will improve the ability of ST to provide insight into patient outcomes before, during, and after behavioral care. Once validity and reliability are established, future work can begin to establish instrumental correlates and meaningful clinical changes within each MSP-V domain. Over time, this work could lead to the creation of cut-off scores for individual domains to guide recommendations for behavioral vs. medical treatment. Future work will also examine how patient-related factors (e.g., age, gender, socioeconomic status, personality, healthcare literacy, etc.) interact with MSP-V domains, further guiding prognosis and treatment recommendations. The work therefore represents a necessary first step in a line of research that can enhance how ST is used to make clinical decisions and to individualize voice therapy.

To evaluate the validity of the MSP-V, the following research questions (RQ) were examined. RQ 1 examined the scale’s content validity, RQ 2 examined construct validity, RQ3 examined concurrent validity, and RQs 4 and 5 examined predictive validity.

1. Do practicing voice-specialized SLPs judge the current domains to be relevant and properly worded?
2. Do the current domains measure the same underlying construct (i.e., stimulability testing), as measured by an Exploratory Factor Analysis?
3. Do the proposed domains capture differences between categorical ratings of stimulability level (e.g., “moderately stimulable” vs “mildly stimulable”) across diagnostic groups?
4. Do the proposed domains differ between diagnostic groups (functional, structural, neurological)?
5. Do the proposed domains predict which patients will return for voice therapy and report practice between assessment and treatment?

## Methods

### Development of the MSP-V

#### Conceptualization

Previous work (Coleman et al., 2025; Toles & Young, 2023; Young, 2025, 2026), as well as clinical experience, suggested that practicing voice-specialized SLPs considered multiple domains when determining the stimulability level of a person or behavior. Thus, providing a single-domain ST outcome scale would likely be strongly influenced by the underlying domains assessed by individual raters. The MSP-V was therefore conceptualized as a multidomain scale, where domains could be measured regardless of a patient’s presentation and the evaluating clinician’s approach to ST with said patient.

When selecting domains including in the MSP-V, two different approaches were considered. One option was to conduct a formalized Delphi consensus protocol using the twenty-seven domains generated in Young (2025). This traditional psychometric “top-down” approach, while time-intensive, would lead to the generation of moderately large pool of domains (e.g., 10-15) that could then be systematically evaluated and winnowed down until a clinically feasible number of domains (e.g., 3 - 5) was achieved. An alternate option was to utilize a “ground-up” approach, where the initial domains were chosen based on discussions with stakeholders grounded primarily on clinical significance and feasibility. Like the top-down approach, several rounds of data collection would be used to systematically evaluate each domain, but with the purpose of adding, replacing, or modifying domains rather than removing extraneous domains. Given the clinical focus of ST, the authors opted for a ground-up approach for the item generation for the MSP-V.

Domains considered for inclusion in the MSP-V were generated primarily based on the results of Young (2025), although information gathered from a review of the existing literature and discussion with interested practicing voice-specialized SLPs around the country was also considered. The generated list of potential domains was broadly categorized into those describing patient psychosocial characteristics (e.g., motivation to attend therapy, openness to therapeutic experience), patient sensory characteristics (e.g., awareness of internal sensations), and patient capacity for behavioral change (e.g., ability to mimic clinician models, responsiveness to cueing). ST is generally defined as the evaluation of an individual’s ability to modify a behavior using therapeutic techniques (Gartner-Schmidt & Gillespie, 2021, 2021; Litts et al., 2015; Toles & Young, 2023; Young, 2025, 2026); thus, the core purpose of ST is to assess capacity for behavioral change. Domains that described patient capacity for behavioral change were therefore prioritized over other domains. While psychosocial and sensory characteristics undoubtedly influence ST outcomes (e.g., Toles et al., 2024), they are not a core element of ST per existing definitions. Additionally, such outcomes can be assessed using existing, validated measures (e.g., the Interoception in Voice Scale [IVP]; Jones et al., 2026; the University of Rhode Island Change Assessment of Voice [URICA-Voice]; Teixeira et al., 2013). Thus, the overarching goal of the MSP-V was to evaluate patient capacity for behavioral change.

An additional goal was to evaluate both clinician and patient perspectives of behavioral change, given the literature suggesting that patient-reported outcomes of ST can influence therapeutic outcomes (Shelly et al., 2023; Toles et al., 2024). Given these goals, three clinician-rated domains that described patient capacity for behavioral change were selected: Degree of Change (DC), Rapidity of Change (RC), and Generalization (G). These domains were selected based on the following rationale. First, these domains represented the concepts frequently endorsed by the SLPs interviewed in Young (2025). Second, each conceptually aligned with distinct concepts within the motor learning literature, namely degree of learning, rate of learning, and transfer of learning (Gentile, 2000; Ranganathan et al., 2022). An additional omnibus domain – Stimulability Level (SL) – was also selected to capture the SLP’s overall impression of patient performance, similar to overall severity ratings in auditory-perceptual evaluations of voice quality (Kempster et al., 2009). This domain was included not only to serve as a familiar anchoring point for clinicians, but also to provide a method of gauging the relationships between each domain and individual clinician’s conceptualization of stimulability level.

The two patient-rated domains, Patient-Rated Voice Quality (PQ) and Patient-Rated Voice Feel (PF), were selected based on the two most-commonly reported outcomes that clinicians asked patients to assess during ST (Shelly et al., 2023; Young, 2025). Inclusion of patient-rated domains allowed not only for a direct measurement of patient-reported change in symptoms, but also an indirect measurement of patient awareness of behavioral change by comparing clinician measures to patient measures. For instance, if a patient was given high ratings on all domains by the clinician but provided low ratings on their self-perceived change scales, the clinician could assume that patient has low awareness of voice changes.

Each domain was designed so that measurement could be completed using a 100 mm visual analog scale (VAS). This measurement scale type was chosen because of its long history of use in voice evaluation research (e.g., Kempster et al., 2009), and because of its advantages over Likert-type or equal-appearing interval scales for ratings of perceptual behavioral data (e.g., Kreiman et al., 1993, 2007). Additionally, the use of a 100-mm VAS would more readily yield normally distributed, ratio data that is suited to standard statistical approaches without transformation. Finally, VAS scales can be more readily be compared to one another, allowing for easy comparison of the MSP-V to established scales using the same measurement methods (e.g., the CAPE-V; Kempster et al., 2009; Voice Therapy Motivation and Voice Perception Inventory; Toles et al., 2024).

A composite domain was also created by taking the average of all clinician-rated and patient-rated domains. This domain was included to give a single-value summary of the patient’s overall response to ST across both clinician and patient perspectives. While this average would be weighted more heavily to the clinician’s perspective, it was of interest to evaluate whether a single, blended score would provide better predictive validity than scores centered in either patient or clinician perspectives. The MSP-V patient and clinician rating form used for data collection is displayed in Figure 1.

**Figure 1.**
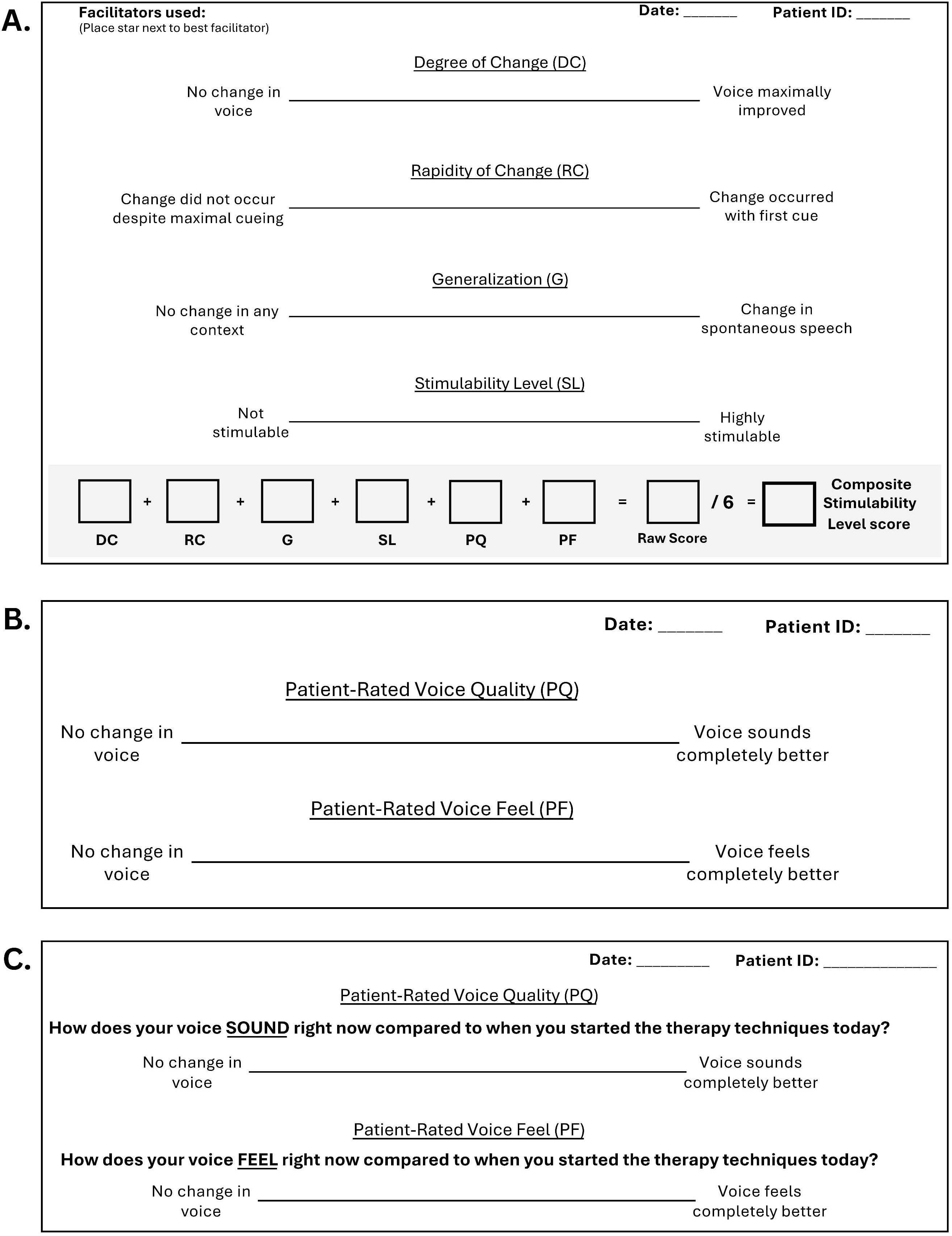
MSP-V Clinician (A) and Patient Rating (B, C) Forms Used for Validation Study. Patient rating form B was replaced with patient rating form C shortly after data collection began to give patients visual access to the instructions for each visual analog scale.

### Data Collection

This study was approved by the UT Southwestern (UTSW) Institutional Review Board (STU-2022-0388). Per IRB protocol, informed consent was waived due to the retrospective nature of the study. Following initial design of the MSP-V, five voice-specialized SLPs at a multidisciplinary voice care center implemented the scale (see Figure 1) into their regular clinical workflow for a six-month period. Each SLP was told the goal of the rating scale (i.e., to quantify the results of ST across in a standardized way across SLPs) and given a general description of each domain. The clinicians were instructed to interpret the scoring of each domain as they saw fit, as no standard rating instructions had yet been developed. SLPs were encouraged to come to the first author with any questions or concerns as they occurred. Minor adjustments were made to the scale throughout the 6 month data collection period, including changing one scale endpoint (the right-most endpoint of the Degree of Change domain from “Voice Normalized” to “Voice Maximally Improved”) four days after initial scale implementation, and placing the instructions to the patients directly on each patient rating scale form rather than having clinicians verbally relay them (see Figure 1C). Patients who were seen for a behavioral voice evaluation at the UTSW Center for Voice Care with a primary voice complaint between Nov 11, 2025 and May 20, 2026, and who underwent ST that was scored using the MSP-V form were included in analysis. Demographic information regarding included participants is provided in Table 1.

**Table 1.** Demographic information for included patients, stratified by diagnosis category.

| Diagnosis | *n* | Age, years<br>*M* (SD) | CAPE-V Severity<br>*M* (SD) | Women<br>*n* (%) | Men<br>*n* (%) |
| --- | --- | --- | --- | --- | --- |
| Functional | 24 | 47.8 (14.5) | 27.0 (18.5) | 18 (75.0) | 6 (25.0) |
| Neurological | 39 | 64.5 (16.5) | 44.2 (19.8) | 18 (53.8) | 21 (46.2) |
| Structural | 15 | 50.3 (18.1) | 32.0 (15.1) | 10 (33.3) | 5 (66.7) |
| Combined Groups | 78 | 56.6 (17.9) | 36.6 (20.0) | 46 (59.0) | 32 (41.0) |

Chart review was used to gather information about each patient in addition to their MSP-V scores. The patient’s voice diagnosis was determined by the referring physician, most commonly one of three fellowship-trained laryngologists at the UTSW Center for Voice Care. Patient diagnoses were categorized as either functional, structural, or neurological for analysis purposes; categorization is presented in Table 2. Each patient’s initial CAPE-V scores (Kempster et al., 2009) provided by their evaluating speech-language pathologist were gathered, as well as whether voice therapy was recommended following ST. Additionally, the patient’s categorical stimulability level classification (i.e., “minimally stimulable”, “stimulable”, “highly stimulable”) was extracted from assessment section of the evaluation note. These categorical classifications of stimulability level have historically been used at this clinical site as an overall rating of the patient’s response to ST and has successfully been used to describe and categorize patients in previous work (McDowell et al., 2023; Turner & Toles, 2025).

**Table 2.** Breakdown of diagnosis categories by laryngologist diagnosis.

| Diagnosis classification (*n*) | Original diagnosis (*n*) |
| --- | --- |
| Structural (15) | Vocal fold scarring (1)<br>Vocal fold cyst (2)<br>Pre-nodular edema/vocal fold nodules (4)<br>Vocal fold polyp/polypoid fullness (4)<br>Chronic laryngitis (1)<br>Vocal fold edema (1)<br>History of anterior glottic squamous cell carcinoma (1)<br>Laryngeal deconditioning s/p oropharyngeal surgery (1) |
| Functional (24) | Muscle Tension Dysphonia (9)<br>Supraglottic compression (9)<br>Voice instability (1)<br>Dysphonia (2)<br>Vocal strain/fatigue (1)<br>Functional dysphonia (1)<br>Transgender woman seeking gender affirming care (1) |
| Neurological (39) | Unilateral vocal fold paralysis (11)<br>Unilateral vocal fold paresis (4)<br>Superior laryngeal nerve palsy (1)<br>Presbyphonia (5)<br>Vocal fold atrophy (9)<br>Glottic insufficiency (2)<br>Vocal tremor (3)<br>Laryngeal dysphonia (1)<br>Parkinson's disease (1)<br>Amyotrophic lateral sclerosis (1)<br>Bilateral paresis (1) |

Initial return to voice therapy was evaluated for patients who were recommended voice therapy during their voice evaluation. Patients were considered to have retuned for voice therapy if they scheduled and completed at least one follow-up appointment during a three-month period after their initial evaluation. A binary measure of documented patient report of practice in the first follow-up appointment was also collected, which was marked as a “yes” if the patient reported any amount of practice between the evaluation and follow up.

### Statistical Analyses

All data analyses were conducted in R version 4.5.3 using the packages *psych* (version 2.6.3), *nfactors* (version 2.4.1.2), and *lme4* (version 2.0-1).

#### Content Validity

There are currently no validated methods for measuring ST outcomes; as such, there is no “gold standard” against which the MSP-V can be compared to evaluate content or concurrent validity. Because of this, the clinical opinions of practicing voice-specialized SLPs was used as the de-facto gold standard for measuring concurrent and content validity. A survey was administered to 9 voice-specialized SLPs around the United States to determine the content validity of the MSP-V domains. SLPs had at least five years of full-time experience working with patients with voice disorders (mean = 11.6, min = 5, max = 27, SD = 6.7) and were recruited evenly from the four geographic areas of the country (South [n = 2], West [n = 2], Midwest [n = 2], Northeast [n = 3]). SLP participants reported currently practicing in a variety of settings, mainly academic voice centers (n = 9), outpatient clinics (n = 4), hospitals (n = 2), and private practice (n = 1), and reported a mean of 85% of their current caseload pertained to voice disorders (min = 50, max = 100, SD = 16.4). Additionally, all SLPs reported performing ST at least 75% of the time during their voice evaluations.

The content validity survey contained a visual depiction of the latest iteration of the MSP-V (see Figure 7), as well as a description of the standard rating procedures developed at the conclusion of the data collection period. They were asked to rate each domain using a using a 4-point scale, where 4 = Highly Relevant; 3 = Quite Relevant or Highly Relevant but Needs Rewording; 2 = Somewhat Relevant; and 1 = Not Relevant. The content validity ratio (CVR; (Lawshe, 1975; Wilson et al., 2012) was calculated for each domain as follows: *CVR = (n_e_ – N/2) / (N/2),* where *N* is the total number of raters and *n*_e_ is the number of raters who gave the item a score of 3 or 4 (i.e., who deemed the item to be relevant). The CVR is a common standardized method of determining content validity for multi-item rating scales with values that range from -1 to +1, with 0 indicating half the raters felt the item was relevant (Almanasreh et al., 2019). Wilson et al. (2012) provides critical values for determining valid scale items based on the number of raters; a cut off of .548 was used to determine domain validity based on the critical values for 9 raters. The Content Validity Index (CVI) was calculated as the mean of the CVR for multiple domains across dimensions of interest (e.g., clinician-rated domains versus patient-rated domains). SLP participants were also asked to describe relevant domains that may have been omitted from the MSP-V in its current version. These answers were analyzed using iterative thematic analysis (Braun & Clarke, 2006).

#### Construct Validity

An exploratory factor analysis (EFA) was conducted to examine the construct validity of the proposed MSP-V domains. Factor analyses are used to assemble large datasets by their underlying “latent” variables. While frequently used in scale development to reduce or eliminate unrelated or trivial items, they can also be used to place items within meaningful categories (Yong & Pearce, 2013). Because the underlying constructs of ST are poorly defined within the literature, an EFA was conducted to test the assumption that the six MSP-V domains describe a similar underlying latent variable – i.e., that of “stimulability testing”. The presence of a single primary factor during the EFA would confirm this assumption; the emergence of multiple factors, however, would indicate that multiple latent variables are present within the MSP-V. The Composite domain was excluded from the EFA as it would be unevenly weighted towards clinician-perceived domains.

The EFA was conducted using a promax (oblique) rotation due to the assumption that factors were likely correlated. Factor extraction was determined using a combination of Kaiser’s criterion (eigenvalue > 1; Kaiser, 1960) and visual examination of scree-plots, as has been suggested in the literature to minimize the risk of overestimation the number of factors due to the examination of eigenvalues alone (Jones et al., 2026; Yong & Pearce, 2013).

#### Concurrent Validity

Concurrent validity was assessed by comparing the documentation-standardized categorical metrics of stimulability level reported within the assessment section of the patient’s evaluation note (i.e., “minimally stimulable”, “somewhat stimulable”, “stimulable”, “highly stimulable”) to each of the MSP-V domains. A series of linear models were conducted, with each MSP-V domain acting as the dependent variable and the categorical stimulability level ratings entered as the independent variable. For each model, the “stimulable” rating was set as the reference value.

#### Predictive Validity

Given the reported importance of ST for informing differential diagnosis and prognosis for behavioral therapy (Coleman et al., 2025; Sund et al., 2021; Toles & Young, 2023), the predictive ability of the MSP-V domains for these were also evaluated. A series of linear models was conducted to assess if any of the MSP-V domains successfully differentiated between the three diagnostic groups (functional, structural, and neurological). Linear mixed-effects models were attempted with a random intercept of clinician, but these models either failed to converge or yielded extremely small ICCs (i.e., 0.06 or less), indicating that the variance between clinicians was relatively small and did not support the use of random modeling. For each linear model, the MSP-V domain was entered as the dependent variable and the categorical diagnostic group was entered as the independent variable.

A series of logistic regressions were conducted to examine the effect of MSP-V domains on both patient initial return for therapy and documented patient-reported practice between evaluation and therapy. For return for therapy, each MSP-V domain was entered as the independent variable and return for therapy (binary yes/no) was entered as the dependent variable. A similar structure was used for the models examining the effect of each MSP-V domain on reported practice. Only patients who were recommended for therapy and who did not receive surgical intervention were included in the analyses for therapeutic follow up and practice, as stimulability ratings could reasonably be expected to change after such surgical procedures. Results were converted into odds ratios for interpretation.

## Results

### Content Validity

The nine SLPs unanimously agreed that the Degree of Change domain and the two patient-rated domains were highly relevant to the theoretical concept of stimulability testing (See Table 3). The Stimulability Level and Generalization domains likewise received a content validity ratio (CVR) value that exceeded the critical value for content validity (Wilson et al., 2012), indicating these domains are relevant when measuring ST outcomes. However, the Rapidity of Change domain did not exceed the critical value, indicating disagreement among the expert raters regarding the necessity of this concept when measuring ST outcomes. The Content Validity Index (CVI) exceeded the critical value when calculated across clinician-rated domains, patient-rated domains, and both clinician-rated and patient-rated domains.

**Table 3.** Content Validity Ratio (CVR) and Content Validity Index (CVI) scores for individual domains and domains separated by rater groups.

| Domain | CVR |
| --- | --- |
| Degree of Change | 1.00* |
| Rapidity of Change | -0.11 |
| Generalization | 0.56* |
| Stimulability Level | 0.78* |
| Patient-Rated Voice Quality | 1.00* |
| Patient-Rated Voice Feel | 1.00* |
|  | CVI |
| Clinician-Rated Domains | 0.56* |
| Patient-Rated Domains | 1.00* |
| All Domains | 0.70* |

\* = above critical value of 0.548 for adequate content validity provided by Wilson et al., (2012) for nine raters

### Construct Validity

Based on Kaiser’s criterion (eigenvalues > 1) and visual analysis of the scree plot, a two-factor model was retained. Together, these two factors accounted for 71% of the variance in the data. As shown in Table 4, the factor loading pattern suggests two latent variables within the data; one primarily explaining variability within the clinician-rated domains and the other primarily explaining variability within the patient-rated domains. The correlation between factors was 0.38, justifying the use of an orthogonal approach. Together, these findings suggest that the scale captures two related but distinct dimensions, reflecting clinician and patient perspectives.

**Table 4.** Exploratory Factor Analysis Factor Loadings.

| Domain | Factor 1 | Factor 2 |
| --- | --- | --- |
| Degree of Change | 0.61 | 0.10 |
| Rapidity of Change | 0.79 | -0.08 |
| Generalization | 0.77 | -0.03 |
| Stimulability Level | 1.00 | 0.00 |
| Patient-Rated Voice Quality | 0.01 | 1.00 |
| Patient-Rated Voice Feel | -0.02 | 0.65 |

### Concurrent Validity

Comparisons for the categorical ratings of stimulability level are displayed in Figure 2 and Table 5. There was a generally descending pattern in all MSP-V domains as patients moved from “highly stimulable” down to the “minimally stimulable” classification, although the statistical significance of these relationships varied. The difference between the “stimulable” classification and all other classifications was statistically significant for the Generalization, Stimulability Level, and Composite domains, all with large effect sizes (Generalization η² = 0.41, 95% CI = 0.26, 1.00; Stimulability Level η² = 0.62, 95% CI = 0.58, 1.00; Composite η² = 0.58, 95% CI = 0.45, 1.00).

**Figure 2.**
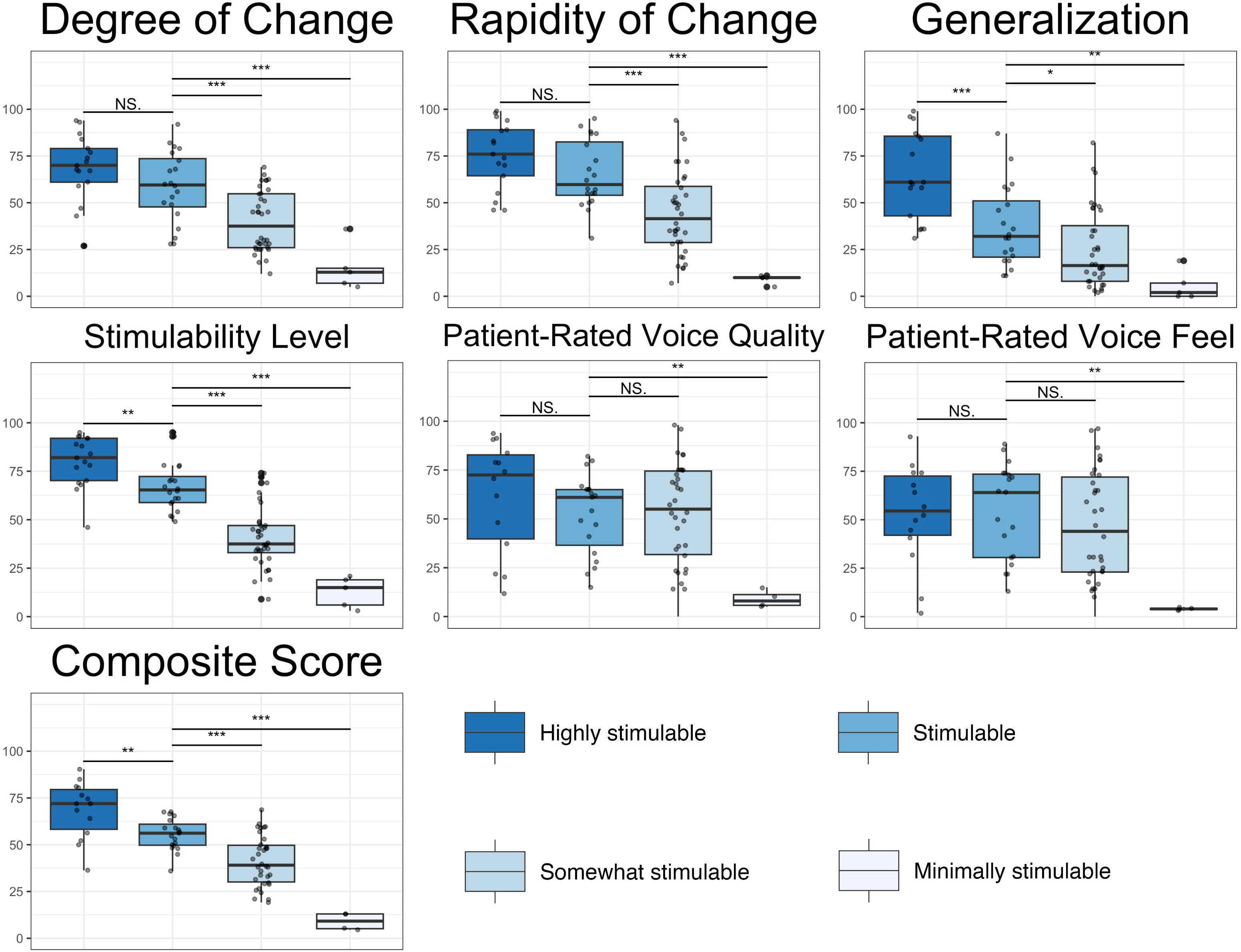
MSP-V Domain scores by Categorical Stimulability Level

**Table 5.** Results of linear regression examining categorical stimulability level ratings for each MSP-V domain.

| **MSP-V Domain** | **Comparison\*** | **B (SE)** | **t (DF)** | **p** |
| --- | --- | --- | --- | --- |
| Degree of Change | Highly Stimulable | 10.17 (5.65) | 1.80 (74) | 0.076 |
|  | Somewhat Stimulable | -18.63 (4.78) | -3.90 (74) | **<.001** |
|  | Minimally Stimulable | -43.40 (8.56) | -5.07 (74) | **<.001** |
| Rapidity of Change | Highly Stimulable | 10.27 (6.47) | 1.59 (74) | 0.117 |
|  | Somewhat Stimulable | -20.87 (5.47) | -3.82 (74) | **<.001** |
|  | Minimally Stimulable | -55.96 (9.81) | -5.71 (74) | **<.001** |
| Generalization | Highly Stimulable | 27.57 (7.00) | 3.94 (74) | **<.001** |
|  | Somewhat Stimulable | -12.53 (5.92) | -2.12 (74) | **0.038** |
|  | Minimally Stimulable | -31.64 (10.61) | -2.98 (74) | **0.004** |
| Stimulability Level | Highly Stimulable | 13.12 (4.54) | 2.89 (74) | **0.005** |
|  | Somewhat Stimulable | -26.46 (3.84) | -6.89 (74) | **<.001** |
|  | Minimally Stimulable | -54.22 (6.88) | -7.88 (74) | **<.001** |
| Patient-Reported Voice Quality | Highly Stimulable | 9.03 (8.68) | 1.04 (74) | 0.302 |
|  | Somewhat Stimulable | 0.11 (7.06) | 0.02 (74) | 0.988 |
|  | Minimally Stimulable | -43.68 (13.56) | -3.22 (74) | **0.002** |
| Patient-Reported Voice Feel | Highly Stimulable | -1.48 (9.23) | -0.16 (74) | 0.873 |
|  | Somewhat Stimulable | -7.47 (7.51) | -1.00 (74) | 0.323 |
|  | Minimally Stimulable | -50.26 (14.42) | -3.49 (74) | **0.001** |
| Composite Score | Highly Stimulable | 13.02 (4.38) | 2.97 (74) | **0.004** |
|  | Somewhat Stimulable | -14.28 (3.56) | -4.01 (74) | **<.001** |
|  | Minimally Stimulable | -46.54 (6.84) | -6.80 (74) | **<.001** |

Notes: *p* values of <.005 are presented in bold text. \*All comparisons were against the reference category of “Stimulable”.

### Predictive Validity

#### Diagnosis Category

The differences in diagnostic category as measured by each MSP-V domain are outlined in Table 6 and Figure 3. No MSP-V domains significantly differentiated among diagnostic categories. However, visual trends appear for certain domains; for example, patients with structural voice diagnoses scored higher, on average, in the Rapidity of Change and Generalization domains than patients with either functional or neurological diagnoses.

**Figure 3.**
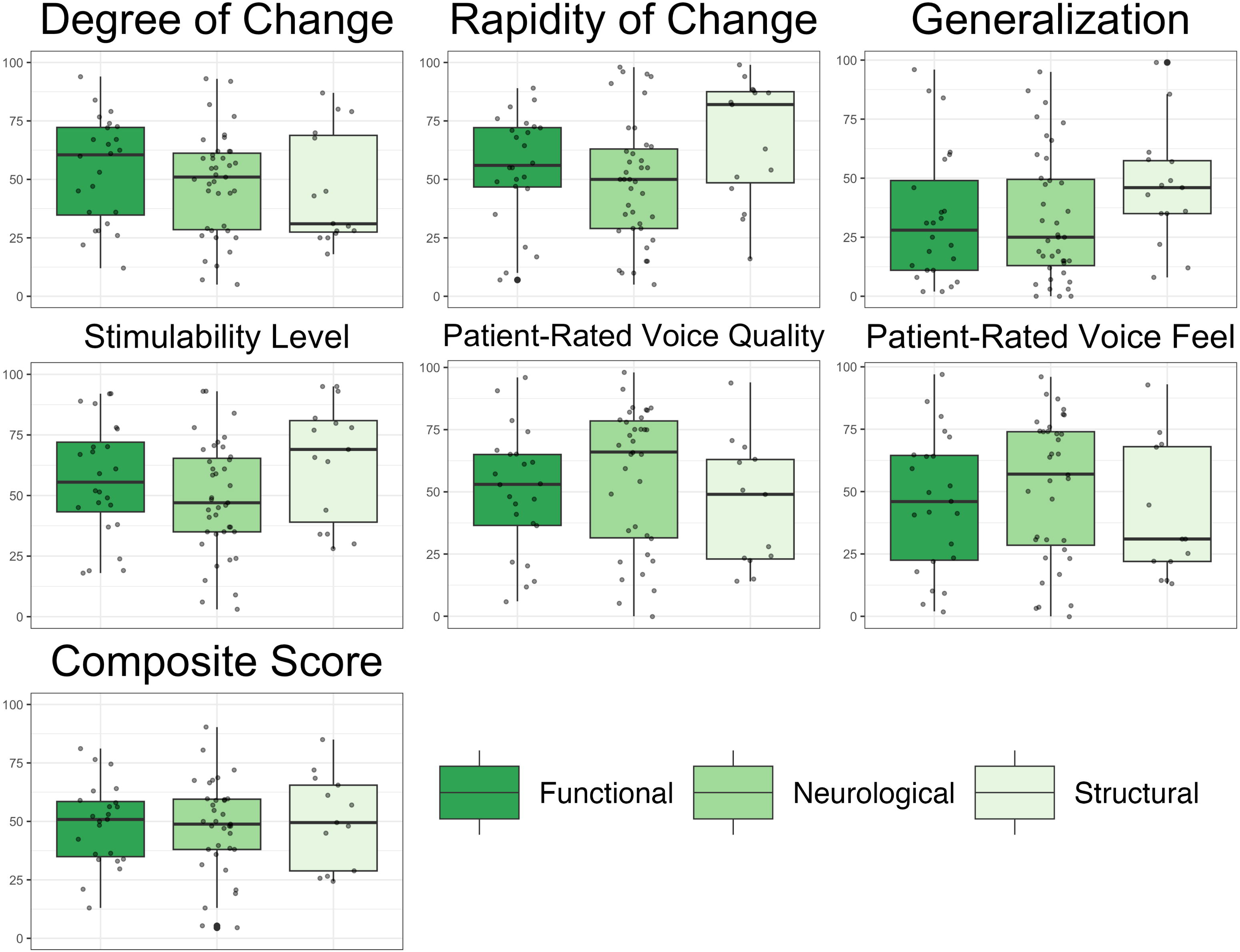
MSP-V Domain scores by Diagnosis Category

**Table 6.** Results of linear regression examining differences in each MSP-V domain by diagnosis category.

| MSP-V Domain | Comparison* | *B* (SE) | *t* (DF) | *p* |
| --- | --- | --- | --- | --- |
| Degree of Change | Neurological | -6.12 (5.84) | -1.05 (75) | 0.298 |
|  | Structural | -8.54 (7.41) | -1.15 (75) | 0.253 |
| Rapidity of Change | Neurological | -6.46 (6.60) | -0.98 (75) | 0.330 |
|  | Structural | 12.03 (8.37) | 1.44 (75) | 0.155 |
| Generalization | Neurological | -0.53 (6.99) | -0.08 (75) | 0.940 |
|  | Structural | 13.02 (8.87) | 1.47 (75) | 0.146 |
| Stimulability Level | Neurological | -7.86 (6.07) | -1.29 (75) | 0.200 |
|  | Structural | 8.03 (7.70) | 1.04 (75) | 0.301 |
| Patient-Reported Voice Quality | Neurological | 5.99 (7.14) | 0.84 (75) | 0.405 |
|  | Structural | -5.12 (9.23) | -0.56 (75) | 0.581 |
| Patient-Reported Voice Feel | Neurological | 5.88 (7.53) | 0.78 (75) | 0.438 |
|  | Structural | -5.62 (9.73) | -0.58 (75) | 0.566 |
| Composite Score | Neurological | -0.99 (5.14) | -0.19 (75) | 0.848 |
|  | Structural | 1.76 (6.64) | 0.27 (75) | 0.791 |

Notes: *p* values of <.005 are presented in bold text. \*All comparisons used the Functional diagnosis category as the reference category

Given the large number of patients with neurological voice diagnoses in this dataset compared to other diagnoses (n = 39, compared to n = 24 functional and n = 15 structural), exploratory analyses were also conducted to determine if the MSP-V domains were sensitive to differences between neurological subclassifications. Patients classified with neurological voice disorders were subdivided into three subcategories: unilateral paralysis (i.e., any patient presenting with unilateral vocal fold paralysis or paresis; *n* =16), atrophy (i.e., any patient presenting with presbyphonia, vocal fold atrophy, or glottic insufficiency due to age-rated changes; *n =* 15), and other (i.e., vocal tremor, laryngeal dystonia, and other miscellaneous diagnoses; *n =* 8). Linear models were conducted to determine if any of the MSP-V domains differentiated between the three neurological subcategories. As illustrated in Figure 4, the Generalization domain distinguished between the neurological subcategories, where patients with unilateral paralysis scored significantly higher than those with atrophy (*B* = 23.40, SE = 9.24, *t*(36) = 2.53, *p* = 0.016). Results of other non-significant models are supplied in the supplemental material.

**Figure 4.**
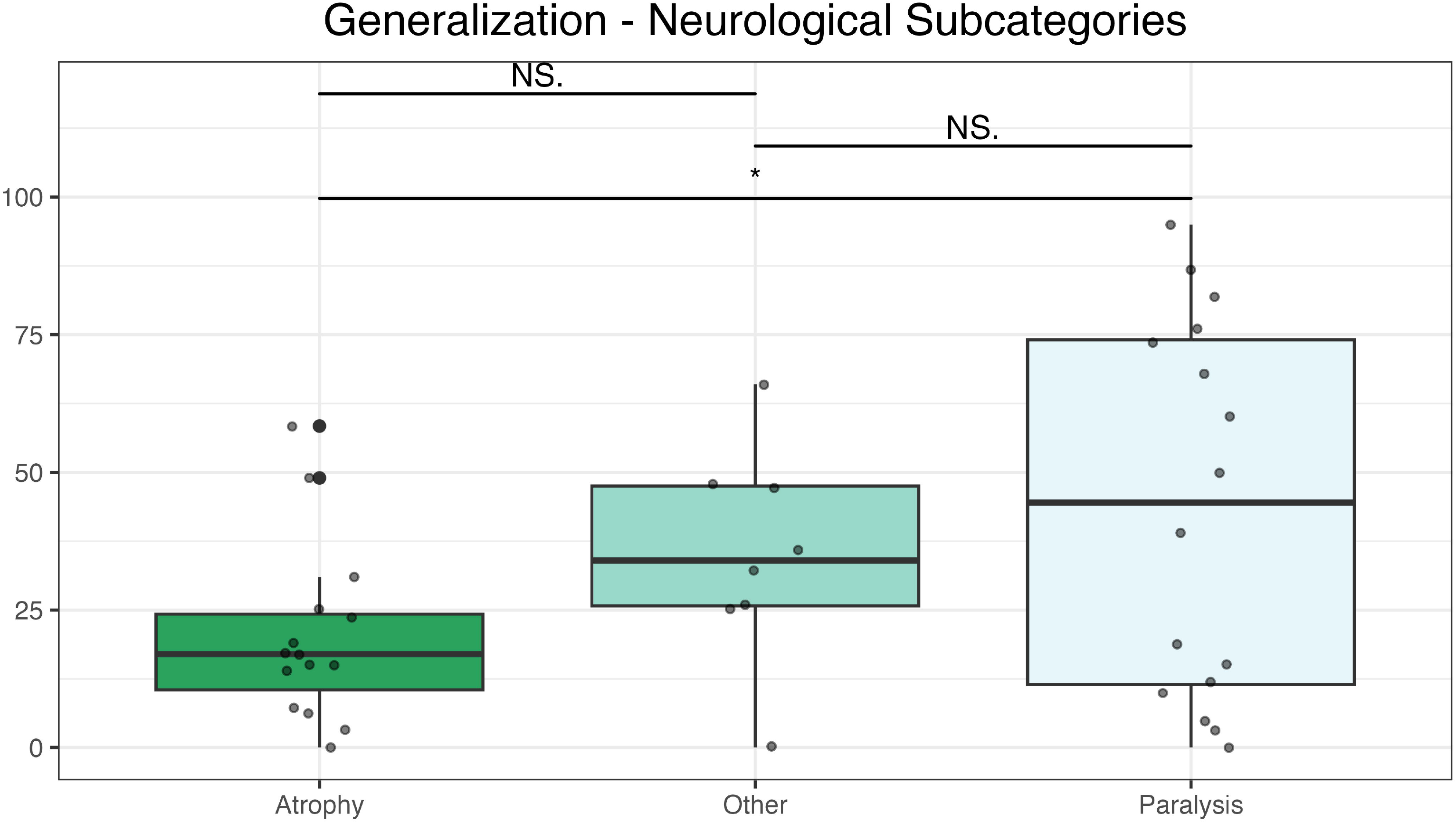
Generalization Domain Scores by Neurological Subcategories

#### Return for Therapy

Of the 78 patients in the database, 66 (85%) patients were recommended for behavioral intervention only, 7 (9%) for both behavioral and surgical intervention, and 1(2%) for surgical intervention only. Only patients who were recommended for behavioral intervention only included for analysis, which resulted in 66 patients available for analysis. Of these patients, one was excluded because they were recommended for voice therapy during their initial evaluation but followed up only for swallowing care. This resulted in a final database of 65 patients for analysis. Forty-two (65%) of these patients returned for at least one voice therapy visit during the three months following their initial evaluation.

The logistic regressions examining each MSP-V domain on the patient’s likelihood of returning to therapy are presented in Table 7 and Figure 5. Two MSP-V domains significantly predicted return for therapy: Rapidity of Change (OR = 0.97, *p* = .016) and Stimulability Level (OR = 0.97, *p* = .013). Within these two domains, patients who were rated more highly on the domain were less likely to return for therapy. In other words, with each point increase of either Rapidity of Change or Stimulability Level, there was an approximately 3% decrease in the odds of the patient returning for therapy. This trend was mirrored, although non-significant, in the other clinician-rated domains. In contrast, the trend was reversed, but also non-significant, in the patient-rated domains, where patients were more likely to return for practice as scores in the patient-rated domains increased.

**Figure 5.**
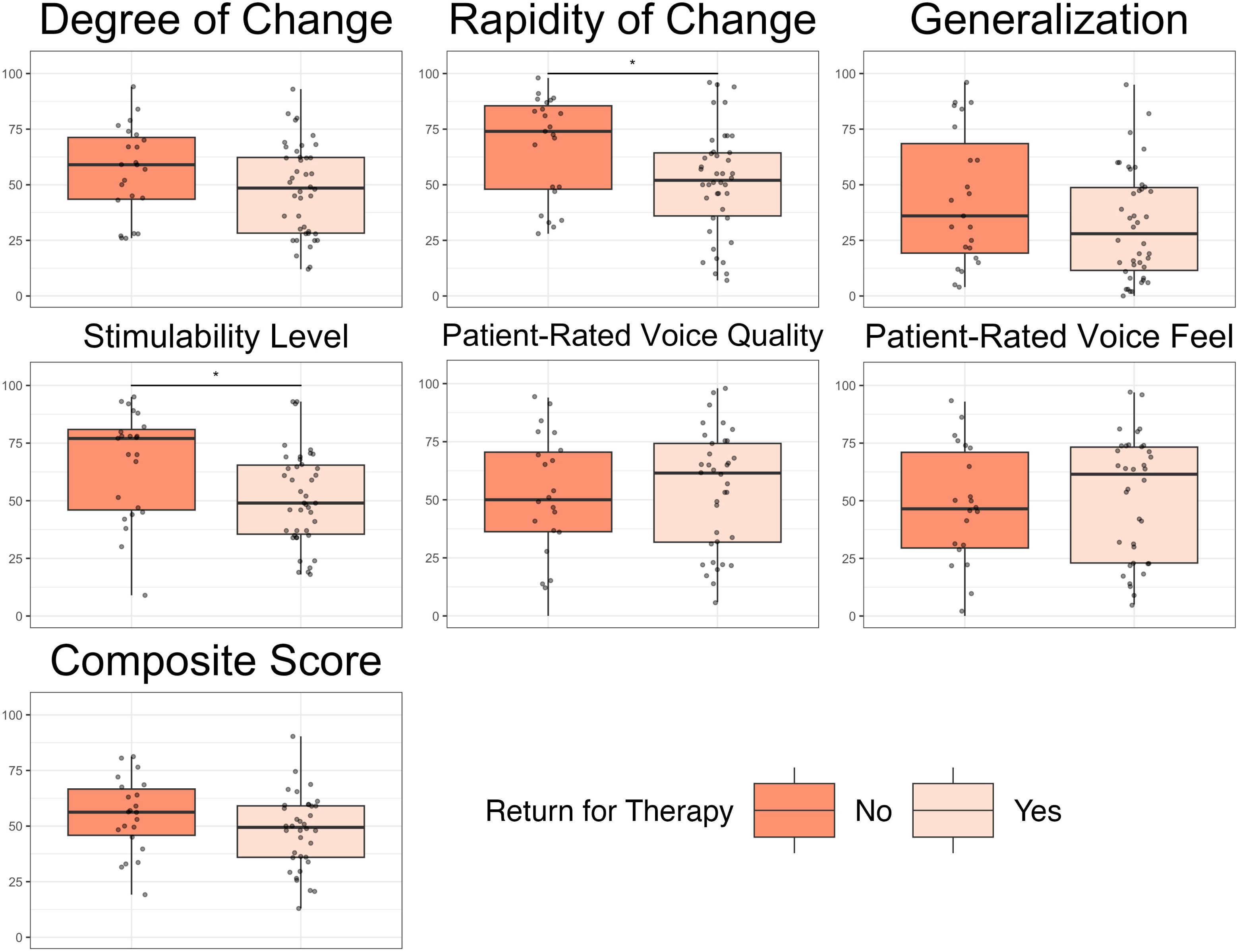
MSP-V Domain Scores by Patient Initial Return for Voice Therapy

**Table 7.** Results of logistic regression examining differences in each MSP-V domain by patient return for voice therapy.

| MSP-V Domain | Est (SE) | OR (95% CI) | *z* (df) | *p* |
| --- | --- | --- | --- | --- |
| Degree of Change | -0.02 (0.01) | 0.98 (0.95 – 1.01) | -1.51 (63) | 0.132 |
| Rapidity of Change | -0.03 (0.01) | 0.97 (0.95 – 0.99) | -2.41 (63) | **0.016** |
| Generalization | -0.02 (0.01) | 0.98 (0.96 – 1.00) | -1.68 (63) | 0.094 |
| Stimulability Level | -0.03 (0.01) | 0.97 (0.94 – 0.99) | -2.48 (63) | **0.013** |
| Patient-Reported Voice Quality | 0.00 (0.01) | 1.00 (0.98 – 1.03) | 0.45 (56) | 0.656 |
| Patient-Reported Voice Feel | 0.01 (0.01) | 1.01 (0.99 – 1.03) | 0.60 (56) | 0.497 |
| Composite Score | -0.03 (0.02) | 0.97 (0.94 – 1.01) | -1.53 (56) | 0.127 |

Notes: *p* values of <.005 are presented in bold text

#### Documented Report of Home Practice

Thirty-two of the 42 patients who returned for therapy (76%) reported practicing voice therapy techniques between their evaluation and their initial return to therapy. None of the MSP-V domains were significant predictors of patient-reported practice (see Table 8 and Figure 6). However, a similar trend in patient-reported practice was noted as was seen in return to therapy; patients who were rated higher on clinician-reported domains were generally less likely practice, while patients who rated themselves higher on patient-reported domains more generally more likely to practice (particularly patients who reported greater changes in voice feel).

**Figure 6.**
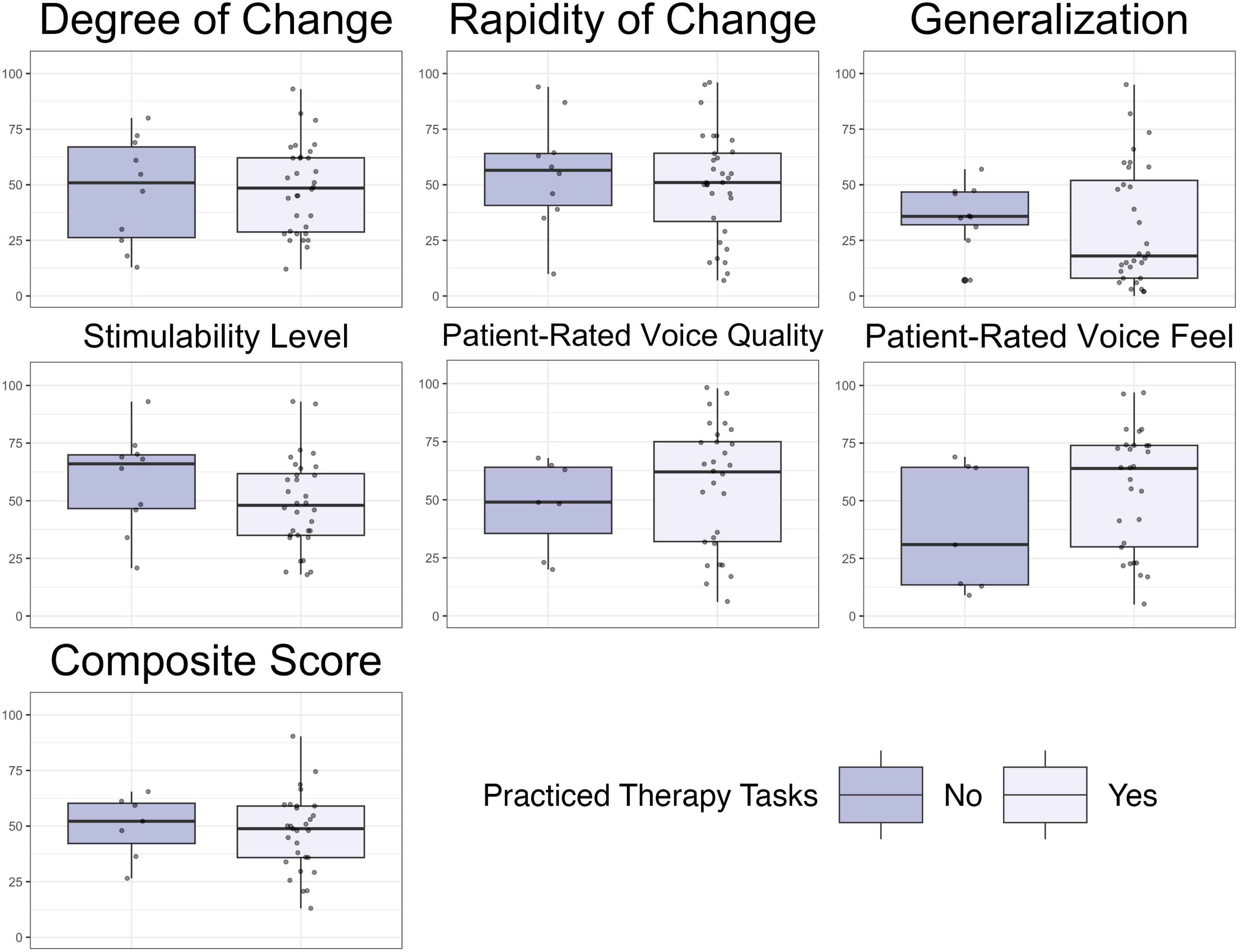
MSP-V Domain Scores by Documented Patient Report of Home Practice between Assessment and Return for Voice Therapy

**Table 8.** Results of logistic regression examining differences in each MSP-V domain by document patient report of practice during first therapy session.

| **MSP-V Domain** | **Est (SE)** | **OR (95% CI)** | **z (df)** | **p** |
| --- | --- | --- | --- | --- |
| Degree of Change | 0.00 (0.02) | 1.00 (0.97, 1.04) | 0.160 (40) | 0.873 |
| Rapidity of Change | -0.01 (0.02) | 0.99 (0.96, 1.02) | -0.61 (40) | 0.540 |
| Generalization | -0.01 (0.01) | 0.99 (0.96, 1.02) | -0.72 (40) | 0.474 |
| Stimulability Level | -0.02 (0.02) | 0.98 (0.94, 1.01) | -1.33 (40) | 0.185 |
| Patient-Reported Voice Quality | 0.01 (0.02) | 1.01 (0.98, 1.05) | 0.74 (34) | 0.459 |
| Patient-Reported Voice Feel | 0.02 (0.02) | 1.03 (0.99, 1.06) | 1.46 (34) | 0.145 |
| Composite Score | -0.01 (0.03) | 0.99 (0.94, 1.04) | -0.38 (34) | 0.701 |

**Table 9.**
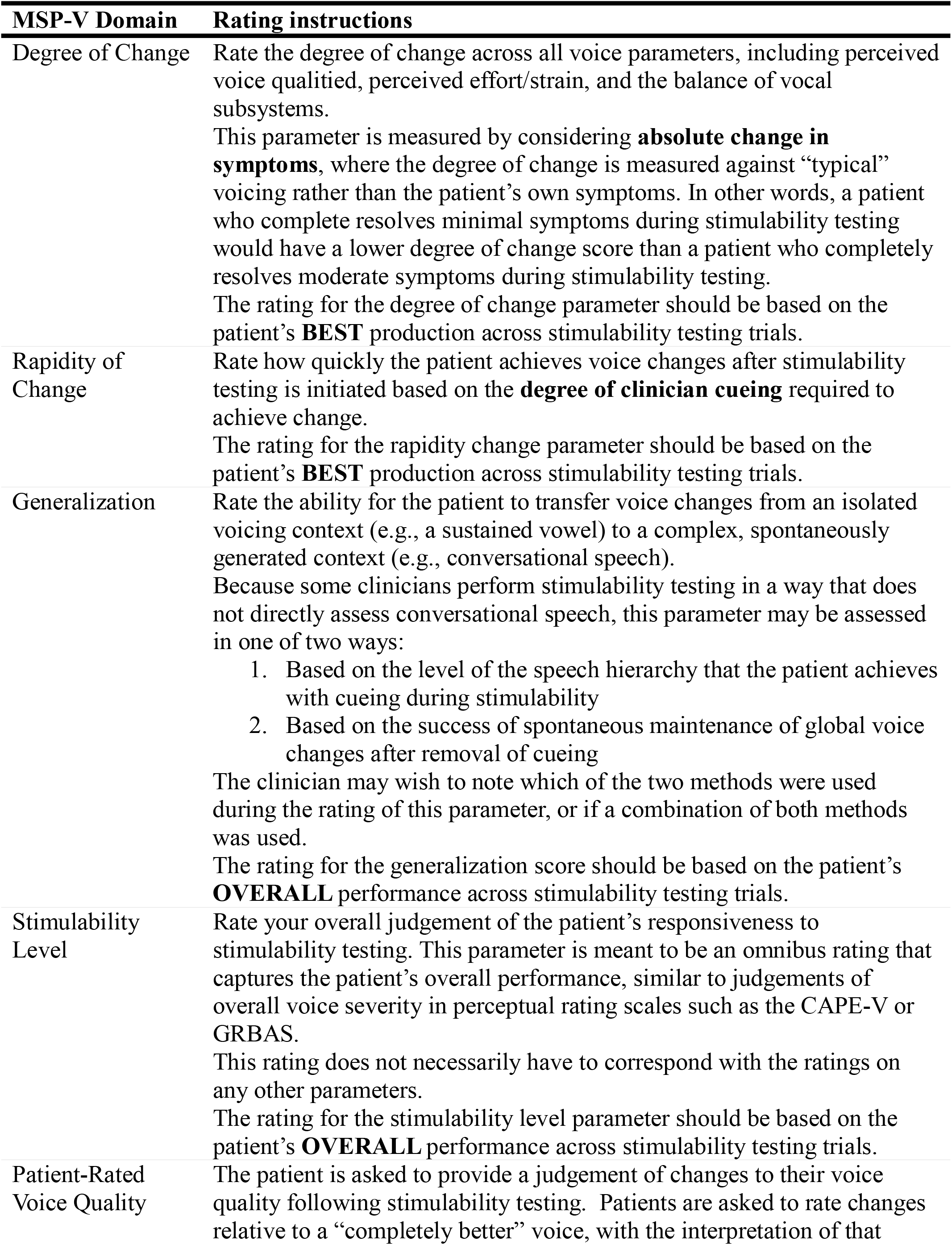
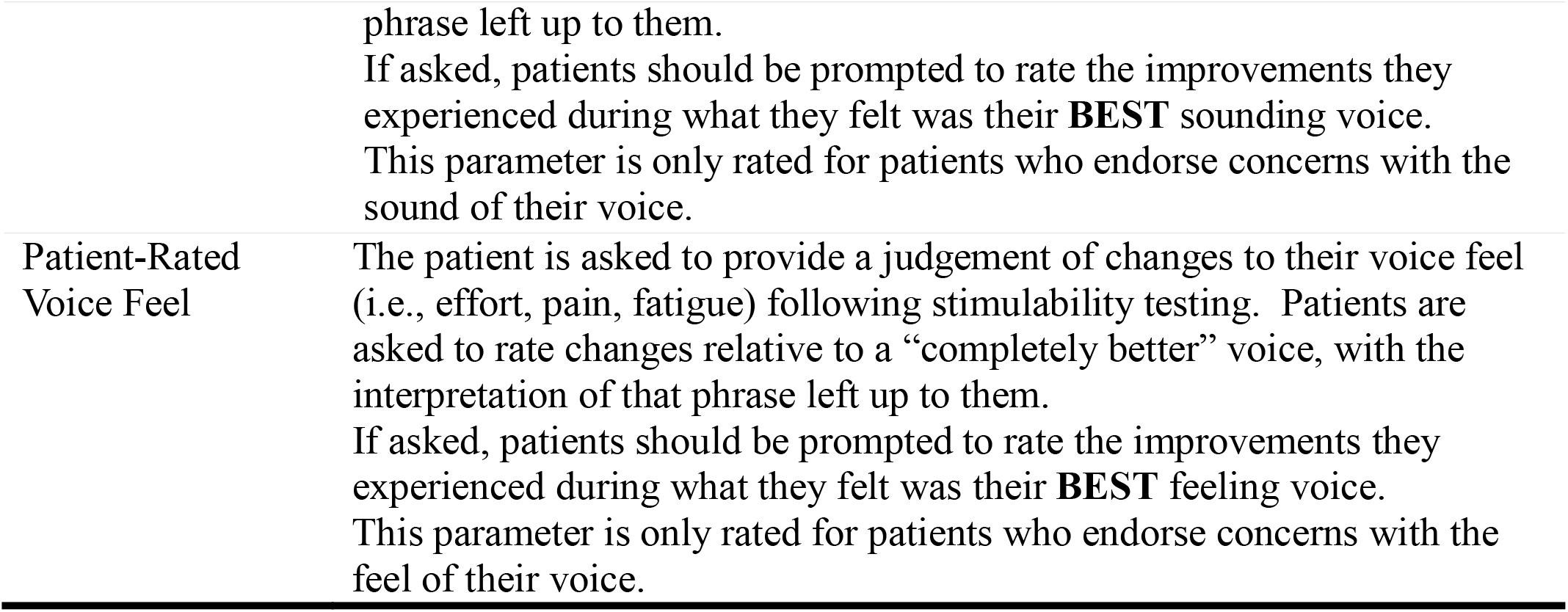
Standardized rating instructions generated following feasibility testing.

## Discussion

The purpose of the current study was to perform preliminary validation for a multi-domain rating scale for stimulability testing (ST) outcomes. In previous studies, ST outcomes are often measured with an omnibus metric of the degree to which a patient or behavior is “stimulable” (e.g., McDowell et al., 2023; Shelly et al., 2023; Weston et al., 2023). However, recent work has shown that many different domains contribute to the formation of a clinician’s judgement of whether the patient or their behavior is considered “stimulable” (Young, 2025, 2026). To address this, the Multidimensional Stimulability Profile for Voice (MSP-V) was designed to measure multiple dimensions potentially relevant to ST outcomes. While preliminary, the results indicate emerging content, construct, concurrent, and predictive validity for the MSP-V. The results of each validity type will be discussed in turn below, as well as the implications of each on future research for ST outcome measurement.

### Content Validity

The content validity for each of the MSP-V domains was assessed by nine voice-specialized SLPs. The Degree of Change, Patient-Reported Voice Quality, and Patient-Reported Voice Feel had the strongest content validity, with all surveyed SLPs agreeing that these domains are relevant to ST. The Stimulability Level domain likewise had relatively high agreement across raters. In contrast, however, two domains, Generalization and Rapidity of Change, had lower agreement across raters, with the Rapidity of Change domain failing to meet the critical value for Lawshe’s Content Validity Index (CVI).

Rather than undermining the utility of a multidomain tool such as the MSP-V, the authors propose that these results highlight the underlying conceptual discrepancies at the core of measuring ST outcomes. While some metrics of patient success during ST appear to have strong conceptual cohesion among the practicing voice-specialized clinicians, other metrics are heavily endorsed by some clinicians but rejected by others. For instance, the Generalization domain was rated as “Highly Relevant” by four SLPs raters and “Not relevant” by one SLP. SLP rater comments on this domain described varying opinions regarding whether skill generalization can and should be assessed during ST, as well as the degree to which generalization is predictive of patient success in therapy. Similarly, the Rapidity of Change domain, which had the greatest disagreement across all domains, did not receive the “Not relevant” rating from any SLP raters. However, many SLP raters commented on factors such as a desire for anchoring points on the VAS for the degree of cueing (e.g., “mild”, “moderate”, “max”) or changes to the scale endpoints. Thus, the poor content validity on these domains is likely partially driven by a combination of internal differences in ST conceptualization and the need for continued refinement of the domain scales themselves for improved clarity and usability. Further work is therefore needed to not only to continue to refine all MSP-V domains but to continue to investigate which domains are most clinically valuable across ST styles. For instance, future work could use qualitative evaluation and standardized clinical samples to determine whether rater disagreement reflects differences in conceptualization of the domains or rather in their operationalization.

Of note, the SLPs participating in the content validity survey were also asked to provide examples of other potential domains of ST that were not currently included within the MSP-V. Although all nine SLPs were asked this question, only one provided a response. This SLP described additional components of ST, including “a patient’s ability to follow directions, unload [supraglottic hyperfunction], [and] access what I am going for during [a stroboscopic] exam”. While these three components are not directly reflected in the MSP-V, each could be captured by existing domains. Change in supraglottic hyperfunction could be assessed using the Degree of Change domain, and patient ability to “follow directions” and “access what I’m going for” could both be considered cues and thus could be scored within the Rapidity of Change domain. The response of raters to solicitation of new MSP-V domains suggest two things. First, the limited response rate suggests that the current MSP-V domains are covering the breadth of domains considered by voice-specialized SLPs when measuring ST outcomes. Second, while the responses that were received could be subsumed within the current MSP-V domains, their presence indicates that the current MSP-V domain descriptions lack adequate depth. As noted above, this indicates that further validation to determine the best methods of describing and measuring each MSP-V domain is required. One potential method for this is to use cognitive interviewing (Willis, 2018) with SLPs who complete the MSP-V to determine factors that influence each MSP-V domain; this is a planned area of future work for continued MSP-V domain development.

### Construct Validity

Exploratory factor analysis was conducted to examine the six MSP-V domains to determine the extent to which they described the underlying construct of “stimulability”. Rather than all domains loading onto a single factor, suggesting a single underlying “stimulability” construct, the domains loaded onto two factors. These results suggest that the four clinician-rated domains measure one construct (i.e., clinician-perceived stimulability), while the two patient-rated domains measure a separate construct (i.e., patient-perceived stimulability). From a psychometric perspective, it is reasonable for ratings performed by separate groups (i.e., patients and clinicians) to make up separate constructs. However, it was interesting that domains that could feasibly measure two separate perceptions of the same construct – namely clinician and patient perception of voice quality change – did not share similar factor loadings or load onto the same factor. This finding underlines a discrepancy between clinician and patient perception of change that occurs during ST, and mirrors that of previous work, where clinicians noted nearly twice the degree of voice quality improvement compared to patients following voice ST (Toles et al., 2024). Additionally, the misalignment between clinician and patient perception of voice change during ST undermines the use of a composite score. While the idea of a single, composite score that represents both clinician and patient response to ST is appealing, results of the EFA suggest it is not supported by the current data. Separate composite scores for clinician- and patient-reported domains may be a suitable alternative, which could be explored in future work.

### Concurrent Validity

The clinicians at the data collection site use a standardized template for their assessment of overall patient ST outcomes. In this template, a drop-down selection allows patients to be categorized as “highly stimulable”, “stimulable”, “somewhat stimulable”, or “minimally stimulable”. Given the lack of a current “gold standard” for ST outcomes, and the fact that the categorical outcomes used within this clinic have successfully been used to describe patients in previous work (McDowell et al., 2023; Turner & Toles, 2025), the MSP-V domains were compared to these categorical classifications of ST as a measure of concurrent validity. It was expected that patients who scored higher on each domain would also be classified into the higher categorical stimulability rating. As seen in Table 5 and Figure 2, this was generally the case for the clinician-rated domains, and particularly so for the Stimulability Level and Composite domains. There was generally a descending pattern in each of the clinician-rated domains as patients were categorized into each descending categorical stimulability rating, indicating that clinician-rated domains captured measurable differences in traditional ST outcomes.

In contrast, this descending pattern was not nearly as strong with the patient-rated domains. Indeed, the only significant contrast for both patient-rated domains was between “stimulable” and “minimally stimulable”. This suggests that the traditional categorical ST outcomes generally do not have a strong linear relationship with patient-reported changes following ST. The results mirror that of the construct validity and indicate ST outcomes measured by clinicians and patients likely represent separate constructs. These findings have important clinical implications. Previous work has varied in whether patient-reported outcomes are considered when describing ST outcomes; some consider solely clinician-rated outcomes (Dejonckere & Lebacq, 2001; Litts et al., 2015; Turner & Toles, 2025), some consider primarily patient-rated outcomes (Shelly et al., 2023), and some consider a mixture of both clinician- and patient-rated outcomes (McDowell et al., 2023; Schneider, 2019; Toles et al., 2024). Importantly, however, all previous work has described both clinician- and patient-reported outcomes using the same language, without considering potential differences in the underlying weighting of each outcome type. The current results suggest that patient-reported ST outcomes may not scale linearly with clinician-reported ST outcomes. Thus, it may be inappropriate to utilize clinician-derived ST outcomes to infer patient-reported ST outcomes. In other words, a patient who is “highly stimulable” from a clinician standpoint may not have necessarily achieved significant gains in their self-perceived voice quality or voice feel. The mismatch between clinician-reported and patient-reported ST is discussed further below.

### Predictive Validity

Preliminary validation of the predictive validity of the MSP-V was performed to determine, (1) the ability of the domains to differentiate between diagnostic categories, (2), the ability of the domains to predict patient return for behavioral therapy, and (3), the ability of the domains to predict patient report of practice. The results suggest emerging predictive validity for both diagnostic differentiation and patient return for therapy in multiple clinician-reported domains.

#### Diagnosis Category

Patients within the current study were categorized by the primary etiology of their voice disorder (functional, structural, or neurological). No significant differences were found across any of the MSP-V domains between each of the voice etiologies. This finding mirrors that of previous work, where categorical stimulability level did not significantly predict diagnosis type (functional, structural, or neurological; McDowell et al., 2023). Several factors likely contributed to this finding. First, there was substantial variability within each domain across each diagnostic category, with patients spanning nearly the entire range of most domains across diagnoses (see Figure 3). Additionally, some diagnostic categories may have been underpowered compared to others; the structural category contained only 15 patients, compared to 39 patients in the neurological category. Given the variability within each diagnostic category seen here, a larger, more balanced sample of patients is needed before determinations can be made regarding the predictive validity of the MSP-V domains for diagnostic differentiation.

Despite this, most of the domains showed visible (albeit not statistically significant) trends that differentiated diagnostic groups from one another. For instance, patients with structural diagnoses on average scored highest on the Rapidity of Change, Generalization, and Stimulability Level domains, but lowest on Degree of Change, Patient-Rated Voice Quality, and Patient-Rated Voice Feel. Patients in the neurological diagnostic category had the opposite pattern, scoring lowest on the clinician-rated domains of Rapidity of Change, Generalization, and Stimulability Level but highest on average in patient-rated domains. Interestingly, patients with functional disorders generally rated between structural and neurological patients, except for Degree of Change, where they had the highest average scores. While these results are preliminary and should not yet be used to draw clinical conclusions, they do suggest that there are differences in how patients with differing etiologies respond to ST. McDowell et al. (2023) similarly reported that patients with functional voice disorders were most likely to be stimulable for voice improvements, followed by structural voice disorders, then neurological voice disorders. However, the same pattern of ST outcomes by etiology found in McDowell et al. (2023) was not replicated in any of the domains of the current study. This may be due to methodological differences between studies, but it could also point to the MSP-V capturing nuance between diagnostic categories that unidimensional ST outcomes do not.

Post-hoc analyses found that the Generalization domain significantly differentiated between patients presenting with vocal fold atrophy and unilateral vocal fold paralysis. Patients with vocal fold atrophy were generally rated quite low on the Generalization domain (mean = 20.00, SD = 16.10), while patents with unilateral vocal fold paralysis (UVFP) were rated more variably (mean = 43.40, SD = 34.12). This is a logical finding within the physiologic context for both diagnoses. Patients with reduced vocal fold muscular mass will likely be able to make short-term gains in vocal quality with cues for increased respiratory drive, loudness, and glottal contact. However, without intervention to increase vocal fold mass via either injection or systematic laryngeal exercise (e.g., PhoRTE; Ziegler et al., 2014), these changes are unlikely to be sustainable beyond short bursts of speech. In contrast, patients UVFP with the paralyzed fold in a relatively medialized position and good vocal fold bulk may be able to make sustainable changes to the voice with facilitative techniques designed to increased vocal fold contact while minimizing compensatory strain.

Thus, when comparing domains across broad diagnostic categories (i.e., structural, functional, and neurological) non-significant trends emerge; however, comparing more specific diagnostic groups resulted in a significant difference within the Generalization domain. Together, these findings suggest that the MSP-V domains may be predictive of specific rather than broad diagnostic category. Future work may benefit from categorizing patients into more specific diagnostic categories to determine if the patterns seen in the current study continue to hold. Should the MSP-V prove useful in differentiating between specific diagnostic categories, future studies could establish clinical cut-offs within each domain for use in the differential diagnosis process for disorders where ST is traditionally used as part of the assessment battery (e.g., muscle tension dysphonia vs laryngeal dystonia; Roy et al., 2007).

#### Return for Therapy

Two clinician-rated domains, Rapidity of Change and Stimulability Level, significantly predicted patient return for therapy within the three months of their initial voice evaluation. This study is the first to find a relationship between voice ST outcomes and patient return for therapy; two previous studies have found no relationship between patient ST level and return for therapy (McDowell et al., 2023; Weston et al., 2023). Interestingly, the patients who returned for therapy were those who were scored significantly lower on each of these domains. This finding is somewhat counter-intuitive, as one would assume that patients who are “more stimulable” would be more invested in therapy and therefore more likely to return. Indeed, previous work has found that patients’ motivation to participant in therapy does increase following ST, and those that are more motivated are more likely to return for therapy (Toles et al., 2024). However, that study examined the relationship between motivation and performing ST, not ST outcomes; thus, performing ST, regardless of the outcome, may increase patient motivation over and above not performing ST.

It is also interesting to note that while patients who scored lower on all clinician-reported domains were more likely to return to therapy, patients who scored themselves higher on patient-reported outcomes tended to be *more* likely to return. This pattern reversal lends further support to the conceptual separation between “clinician-perceived” stimulability and “patient-perceived” stimulability. Further, it provides important insight into in the potential driving factors behind patient motivation to pursue behavioral voice care following ST. Based on the patterns found here, it appears that patients who perceive greater improvement are more likely to return, while patients with greater perceived change by the clinicians are less likely to return. It is possible that patients who received higher scores on the clinician domains felt more confident in their ability to improve their voice without clinician guidance and thus did not follow up with therapy, while those with lower scores felt they would benefit from increased clinical feedback. Of course, many factors may contribute to patient return to therapy that are beyond the scope of the current study; based on these findings, however, continued examination into the relative importance of clinician-rated vs patient-rated ST domains into patient return for therapy is a potential avenue for future investigation.

#### Documented Report of Home Practice

No MSP-V domains significantly predicted documented patient report of home practice in their first therapy session. This may have been due to reduced sample size for this analysis compared to other analyses; only those patients who returned for therapy could be included, leaving only 42 patients available for analysis. Future work with a larger sample, ideally balanced between diagnostic categories, is needed to investigate the relationship between patient report of practice and the MSP-V domains.

### Limitations and Future Directions

Several limitations of this initial validation study must be acknowledged. First, the study was conducted at a single clinical site, which limits the generalizability of the results. Further, although the MSP-V is designed to be used regardless of individual ST style, the results in the current study are biased towards the styles of the SLPs in the current study. This style has been described in previous work (McDowell et al., 2023; Turner & Toles, 2025); briefly, it generally involves trialing many facilitators at the phoneme or syllable level and marking each as either “improved” or “not improved” on a standard clinical template. As such, the ratings provided by the SLPs in the current study may align directly with those of an SLP who uses a ST style using a single primary therapeutic approach (e.g., conversation training therapy, as seen in Shelly et al., 2023). Future work assessing the MSP-V across a larger group of SLPs with a variety of ST styles is needed to confirm the results are generalizable regardless of personal ST approach. Additionally, the MSP-V was implemented by five voice-specialized SLPs, who, as practicing clinicians, implemented the protocol to the best of their ability given the many demands on their time and energy. This resulted in uneven data collection between the SLPs, leading to a sample that was weighted towards SLPs with caseloads that were primarily neurologic or functional voice disorders. The uneven weighting between diagnostic groups may have obscured the ability of the MSP-V domains to differentiate between those groups. Ongoing data collection with a larger, more diagnostically balanced data set is underway to assess whether this is the case.

Given the findings of the current study, two core adjustments were made to the original MSP-V rating forms shown in Figure 1. The proposed rating form for ongoing use and validation is presented in Figure 7. The first change was the removal of the composite score. The evidence suggesting separate underlying constructs for clinician-perceived and patient-perceived stimulability undermines the use of a composite score utilizing both clinician-rated and patient-rated domains. The second change is the addition of a list of examples of voice sound deviances (“raspy, scratchy, hoarse, strained”) and voice feel deviances (“tired, sore, painful, like there’s a lump in your throat”) above each corresponding patient-rated domain, alongside a question asking patients if they have concerns with the way their voice sounds or feels (marked as a binary yes/no response). If the patient responds with a “yes” to either domain, they are prompted to provide their perception of change within this domain during ST using a VAS. This change was implemented due to feedback from the five SLPs using the MSP-V in validation study. These SLPs stated that the most common issue they had with using the MSP-V was communicating with patients on how to fill out the VAS. These problems either stemmed from difficulty understanding how the scales themselves functioned or from patients feeling as though they did not have complaints with either voice sound or voice feel. Difficulty with VAS use is a documented issue in test development (Streiner et al., 2015), and is a potential reason for exploring alternate methods of measuring patient-rated domains. In the interim, a form illustrating how to fill out a VAS for patients is also included within the MSP-V directions (see supplemental materials). Allowing patients to opt-out of providing answers to VAS that they feel to not pertain to them should reduce the potential for spurious answers that may falsely inflate results within a particular domain. Additionally, relationships between patient diagnosis, candidacy, and prognosis could still be examined based on the patient’s binary answers (i.e., whether a patient reported complaints with voice sound or voice feel). These two changes were therefore rooted in a combination of analytic and clinical data.

**Figure 7.**
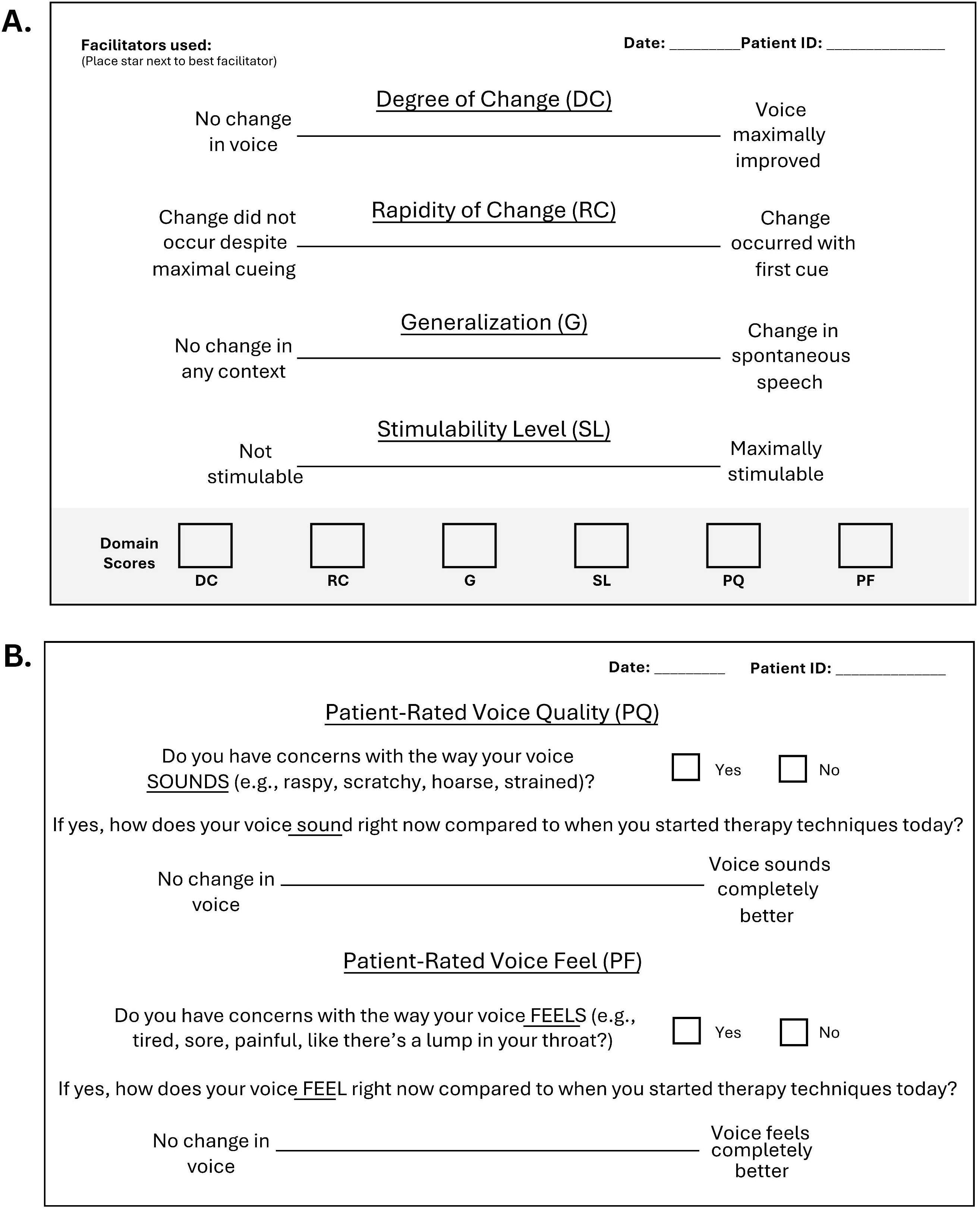
Clinician (A) and Patient (B) Rating Form Used for Content Validation and Suggested for Ongoing Use

Despite the preliminary nature of the current study, the MSP-V shows promise as clinical and research tool that, with continued refinement and further validation, may enhance the diagnostic, prognostic, and therapeutic utility of ST. Areas of planned future refinement and validation include improving content validity of scale items through cognitive interviewing and comparing new scale types and scale wording, examining the construct validity of each domain by comparing each to physiologic changes throughout ST, and examining inter-rater and intra-rater reliability for all domains. After completing planned validation work, future research can begin to expand upon the preliminary predictive validity established in the current study by further examining the relationship between MSP-V domains and patient diagnosis, return for therapy, practice within therapy, and other factors beyond the scope of the current study. Large, multi-site studies using the MSP-V could be utilized to establish normative values and clinically meaningful change within MSP-V domains, which in turn could be used to establish clinical cut-offs used to recommend patients for medical versus behavioral voice intervention. Given the surprising relationship between clinician-reported outcomes and patient return for therapy in the current study, it is of special interest to examine the relationship between patient factors (e.g., age, gender, voice-related handicap, socioeconomic status, personality, etc.), MSP-V domains, and voice care outcomes in future work to further examine how these variables interact. This study thus represents the first step of many towards providing researchers and SLPs a validated tool for quantifying ST results and understanding the relationship between ST and patient outcomes.

## Conclusion

To the authors’ knowledge, there was no validated metric for quantifying the results of stimulability testing (ST) for voice-specialized speech-language pathologists prior to the current study. This study introduces the Multidimensional Stimulability Profile for Voice (MSP-V) as the first method for quantifying multiple outcomes of ST as perceived by both SLPs and patients. The results of this preliminary validation study conducted on 78 patients with voice study indicate emerging content, construct, concurrent, and predictive validity. Construct validation indicated clinician-rated domains consisted of a distinct construct from patient-rated domains. This finding was mirrored in the predictive models for patient return for therapy and patient report of practice within therapy, where clinician-rated domains and patient-rated domains showed contrasting patterns. Despite this, both clinician- and patient-rated domains had good content validity, indicating both aspects are relevant for ST. The results of this validation study indicate a potential role of the MSP-V for clinical and research use in quantifying the results of ST. Ongoing validation of the MSP-V is underway, including collection of inter-rater and intra-rater reliability, validation of domains against physiologic metrics, and ongoing scale refinement.

## Acknowledgements

We gratefully acknowledge the support Melanie Turner Tice, Caroline Hurt, Amy Harris, and Cory Atkinson for their assistance in data collection, as well as their contributions to the refinement of the MSP-V in general. Special thanks to all the speech-language pathologists who participated in the content validity survey for their time and expertise.

## Data Availability Statement

The datasets generated during the present study, as well as the R code used for analysis, are available from the first author upon reasonable request. The MSP-V scale is available for clinical or research use under a CC-BY-ND-SA 4.0 license from the first authors’ website: https://www.elizabeth-d-young.com/

