## Supplemental Files for "Preliminary Validation of a Multidimensional Stimulability Testing Rating Scale for Patients with Voice Disorders"

Table S1. Results of linear regression examining differences in each MSP-V domain by neurological subcategory

| **MSP-V Domain** | **Comparison*** | ***B* (SE)** | ***t* (DF)** | ***p*** |
|---|---|---|---|---|
| Degree of Change | Other | -1.07 (9.82) | -0.11 (32) | 0.914 |
| | Paralysis | -5.05 (8.06 | -0.63 (32) | 0.535 |
| Rapidity of Change | Other | 6.83 (11.81) | 0.58 (32) | 0.566 |
| | Paralysis | 4.99 (9.69) | 0.52 (32) | 0.610 |
| Generalization | Other | 15.05 (11.26) | 1.34 (32) | 0.190 |
| | Paralysis | 23.40 (9.24) | 2.53 (32) | **0.016** |
| Stimulability Level | Other | 1.08 (10.33) | 0.10 (32) | 0.918 |
| | Paralysis | -0.97 (8.48) | -0.11 (32) | 0.910 |
| Patient-Reported Voice Quality | Other | 2.50 (13.64) | 0.18 (32) | 0.856 |
| | Paralysis | 1.06 (11.03) | 0.10 (32) | 0.924 |
| Patient-Reported Voice Feel | Other | -12.66 (13.61) | -0.93 (32) | 0.359 |
| | Paralysis | -11.96 (11.00) | -1.09 (32) | 0.285 |
| Composite Score | Other | 0.43 (9.60) | 0.05 (32) | 0.965 |
| | Paralysis | -0.31 (7.76) | -0.04 (32) | 0.968 |

Notes: $p$ values of <.005 are presented in bold text. *All comparisons used the Atrophy diagnosis subcategory as the reference category

Figure S1. Instruction Page Provided to Clinicians with MSP-V Forms; available under a CC-BY-ND-SA 4.0 license at www.elizabeth.d.young.com

### Instructions

**General instructions:**

1. The clinician form contains four 100 mm visual analog scales (Degree of Change, “DG”, Rapidity of Change, “RC”, Generalization, “G”, and Stimulability Level, “SL”). These scales should be filled out by the clinician immediately upon completion of stimulability testing using a single tick mark upon the 100 mm line. See the MSP-V scoring instructions for additional information regarding how to score each individual domain, including patient-rated domains.

2. The patient form contains two 100 mm visual-analog scales (Patient-Rated Voice Quality, “PQ”, and Patient-Rated Voice Feel, “PF”). These scales should be filled out by the patient immediately upon completion of stimulability testing. If the patient does not experience significant problems with their voice sound or feel, they may opt-out of completing the rating of either domain using the provide check box. Otherwise, written instructions are provided for the patient above each domain. The “How to use patient voice rating scales” page is provided as a guide for patients who express difficulty with visual-analog scale ratings.

3. Once all ratings are complete, use a ruler to determine the score for each of the ratings scales. Place the score for each rating scales in the respective score box at the bottom of the clinician rating form.

   Example: If a clinician gave a patient a degree of change score of 57, they would place the number 57 in the box labeled ”DC” on the bottom of the clinician rating form.

**Miscellaneous:**

4. The clinician and patient rating forms should be stapled together after completion to avoid loss of data

5. There is space at the top of the clinician rating form to indicate the facilitators used during stimulability testing. If desired, a star (or other forms of marking) can be used to indicate which facilitators were most beneficial to the patient

6. The back side of the clinician rating form can be used to take additional notes regarding the results of stimulability testing if desired

7. For best results, print using the “original size” settings to ensure the 100 mm visual-analog lines remain 100 mm during printing.

Figure S2. Instruction Page for Patients to Aid with Visual Analog Scale Completion; available under a CC-BY-ND-SA 4.0 license at www.elizabeth.d.young.com

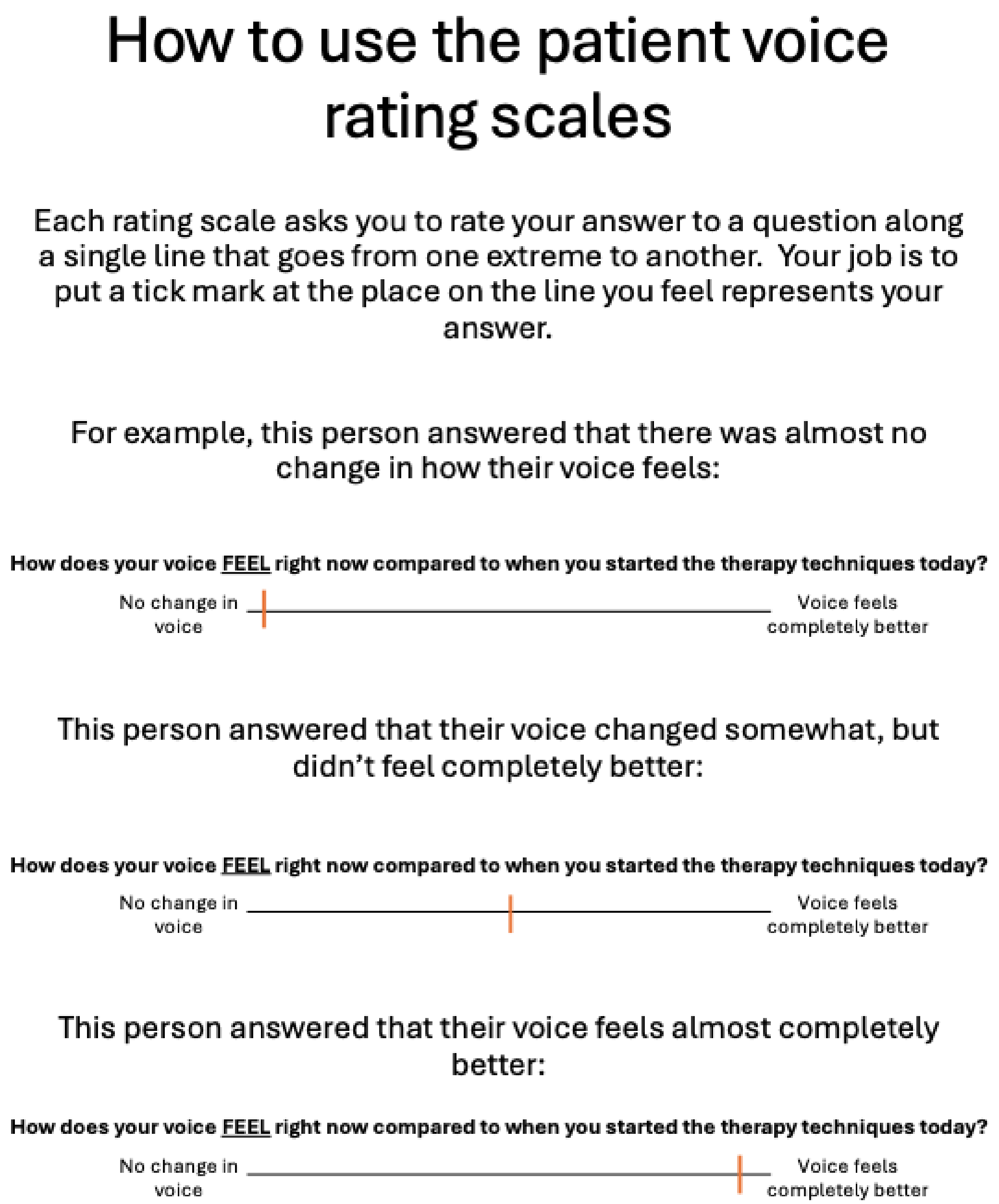

### How to use the patient voice rating scales

Each rating scale asks you to rate your answer to a question along a single line that goes from one extreme to another. Your job is to put a tick mark at the place on the line you feel represents your answer.

For example, this person answered that there was almost no change in how their voice feels:

**How does your voice FEEL right now compared to when you started the therapy techniques today?**

No change in voice | Voice feels completely better

This person answered that their voice changed somewhat, but didn't feel completely better:

**How does your voice FEEL right now compared to when you started the therapy techniques today?**

No change in voice | Voice feels completely better

This person answered that their voice feels almost completely better:

**How does your voice FEEL right now compared to when you started the therapy techniques today?**

No change in voice | Voice feels completely better
